# The Natural History and Burden of Malaria During the First Year of Life in a High Transmission Setting in Uganda

**DOI:** 10.1101/2025.11.17.25340386

**Authors:** Miriam Aguti, Joaniter I. Nankabirwa, Jimmy Kizza, Abel Kakuru, Timothy Ssemukuye, Harriet Adrama, Peter Olwoch, Bishop Opira, Baker Odongo, Kylie Camanag, Miriam Nakalembe, Tamara Clark, Philip J. Rosenthal, Grant Dorsey, Prasanna Jaganathan, Moses R. Kamya

**Affiliations:** Infectious Diseases Research Collaboration, Kampala, Uganda; School of Medicine, Makerere University College of Health Sciences, Kampala, Uganda; Department of Community and Public Health, Busitema University, Tororo, Uganda; Department of Medicine, Stanford University School of Medicine, USA; Department of Medicine, University of California, San Francisco, CA, USA

**Keywords:** Malaria, Infants, Uganda, Parasitemia, Incidence, LLINs, Sickle Cell Trait, Vector Control, Chemoprevention

## Abstract

Infants under one year of age are considered partially protected from malaria due to maternal antibodies and fetal hemoglobin. However, emerging evidence suggests that malaria burden in this age group may be underestimated. We enrolled a cohort of 855 infants in Busia District, Uganda to characterize and identify risk factors for malaria incidence and parasite prevalence during the first year of life. The study was conducted from 2021-2025, prior to malaria vaccine roll-out. Infants born to HIV-uninfected women were enrolled at 4-8 weeks of age and followed 7 days/week in a dedicated study clinic to 1 year of age. Routine visits every 4 weeks included assessments for parasitemia by microscopy and quantitative PCR. Over 706.7 person-years of follow-up, 662 malaria episodes occurred; 71% occurred prior to 9 months of age. Overall prevalence of microscopic parasitemia was 7.9% and combined prevalence of microscopic and submicroscopic parasitemia was 21.8%. Sickle cell trait (HbAS) conferred 39% protection against symptomatic malaria but had no association with risk of parasitemia. Modern housing construction and higher maternal education were independently associated with reduced malaria risk. District-wide distribution of alpha-cypermethrin plus chlorfenapyr long-lasting insecticide-treated nets in October 2023 was followed by an 80% reduction in malaria incidence and significant declines in parasitemia prevalence. These findings underscore the urgent need for age-appropriate preventive interventions targeting young infants, such as earlier vaccine administration and/or monoclonal antibodies, alongside sustained investment in next-generation vector control, and attention to socioeconomic determinants of malaria risk.

## Introduction

Malaria remains a leading public health concern, especially in sub-Saharan Africa, which is responsible for over 95% of global malaria morbidity and mortality^1^. Children under five years of age are disproportionately affected by malaria, and infants under one year represent a vulnerable group that is often understudied ^2, 3^. Although infants younger than six months are often considered partially protected from malaria due to the presence of maternal antibodies^4^ and fetal hemoglobin ^5, 6^, emerging evidence suggests that the burden of malaria in this age group may be substantially underestimated ^2, 7, 8^. Indeed, evidence indicates that infants as young as one to two months of age can experience clinical malaria, with the malaria risk increasing substantially after 2 months of age ^2, 7, 8^.

Malaria prevention strategies targeting infants under one year of age are limited, especially in regions with perennial malaria transmission. In such settings, in addition to LLINs, the World Health Organization (WHO) recommends perennial malaria chemoprevention (PMC). PMC with sulfadoxine-pyrimethamine (SP) following the immunization schedule of the expanded programme on immunizations (formerly referred to as IPTi) has been shown to be safe and effective against malaria in the first year of life,^9^ but uptake has been low, in part because it provides a relatively low level of protection against symptomatic malaria^9^ and its efficacy is limited by growing SP resistance^10^. Recently, two malaria vaccines, RTS,S and R21, have been approved and are beginning to be deployed across Africa, including Uganda, among children 5-17 months of age. However, in prior studies optimal malaria vaccine efficacy was not reached until receipt of 3 doses (at 8-9 months of age), leaving a protection gap for younger infants.

Finally, there are no tailored WHO malaria treatment guidelines for infants under six months of age. The lack of targeted malaria prevention and treatment strategies in very young children, together with the new evidence suggestive of a higher disease burden than previously anticipated in this age-group, highlight an urgent need for research to guide evidence-based policy and clinical practice.

To address these gaps, we conducted a prospective cohort study to characterize the natural history and burden of malaria during the first year of life in a high transmission area of Uganda. We aimed to inform the development of effective, age-appropriate interventions and treatment guidelines for infants living in perennial, high-transmission settings.

## Methods

### Study participants and setting

Infants followed in this cohort study were born to HIV-uninfected women who participated in a double-blind, randomized controlled trial of three different intermittent preventive treatment in pregnancy regimens (DPSP study, NCT04336189) ^11^. Briefly, 2,757 pregnant women were randomly assigned in a 1:1:1 ratio to receive monthly IPTp with either SP, dihydroartemisinin-piperaquine (DP), or a combination of DP and SP, and followed to delivery. Participants were approached at their 4-week post-partum visit and asked if they would like their infants to be enrolled in this follow-up study. Infants were screened and enrolled if they were between 4-8 weeks of age, born to mothers who participated in the DPSP trial, lived in Busia District, had parents/guardians who provided written informed consent, and agreed to come to a dedicated study clinic for all routine medical care and to avoid medications given outside the study clinic. Consecutive enrollment was continued until the desired sample size of 871 infants was achieved.

The study was conducted between November 2021 and January 2025 in Busia District, south-eastern Uganda. Busia is a high malaria endemic district with perennial transmission that is marked by two seasonal peaks. Prior to 2013, vector control in Busia had been limited to targeted distribution of long-lasting insecticide treated nets (LLINs) through antenatal care services. Universal distribution campaigns of free LLINs were conducted in Busia District in May 2013, May 2017, December 2020, and October 2023. The first two campaigns utilized standard pyrethroid LLINs, the 2020 campaign LLINs containing deltamethrin plus piperonyl butoxide (PBO, PermaNet 3.0), and the 2023 campaign LLINs containing alpha-cypermethrin plus chlorfenapyr (Interceptor G2). Indoor residual spraying of insecticides has never been implemented in Busia District.

### Study procedures

At enrollment, a baseline evaluation was conducted on all infants including a detailed medical history, focused physical examination, and blood collection by venipuncture for thick blood smear, quantitative PCR (qPCR), hemoglobin measurement, sickle cell testing, and storage for future molecular studies. Additionally, all infants received a LLIN containing deltamethrin plus piperonyl butoxide (PBO, PermaNet 3.0) at enrollment. Participants with sickle cell disease were given monthly chemoprophylaxis with SP. Study participants were encouraged to come to a dedicated study clinic open 7 days per week for all their medical care and provided with transportation reimbursement. Routine visits were conducted every 4 weeks and included a standardized evaluation and collection of blood by finger prick for thick blood smear, qPCR, hemoglobin measurement (at 8, 24, and 52 weeks), and storage for future studies. Study participants found to have a fever (tympanic temperature *>* 38.0°C) or history of fever in the previous 24 hours at the time of any clinic visit had a thick blood smear read immediately. If the thick blood smear was positive by light microscopy, the patient was diagnosed with symptomatic malaria and managed according to national guidelines^12^. Study participants who missed their scheduled routine visits were visited at home and requested to come to the study clinic as soon as possible. All enrolled participants were followed to one year of age unless they were prematurely withdrawn. Participants were withdrawn if they: 1) moved out of the cohort household; 2) were unable to be located for *>* 3 months; 3) withdrew informed consent; or 4) were unable to comply with the study schedule and procedures.

Study procedures for the mothers have been previously described^13^. Briefly, following enrollment, women were visited at home, where a household survey was conducted to collect detailed information about the participant’s household using a structured questionnaire. Data from this household survey was also applied to infants enrolled in this study.

### Laboratory procedures

Blood smears were stained with 2% Giemsa and read by experienced microscopists. A blood smear was considered negative when the examination of 100 high power fields did not reveal asexual parasites. For quality control, all slides were read by a second microscopist and a third reviewer settled any discrepant readings. Blood samples collected at enrolment and at the time of each routine visit were tested for the presence of sub-microscopic parasitemia using a highly sensitive qPCR assay targeting the multicopy conserved *var* gene acidic terminal sequence with a lower limit of detection of 0.05 parasites/μl^14^. Sickle cell status was ascertained via testing of samples by hemoglobin electrophoresis at the Ugandan Central Public Health Laboratory.

### Data management and statistical analysis

Data was collected using standardized case record forms and entered into Microsoft Access. All statistical analyses were performed using Stata version 19 (StataCorp, College Station, TX, USA).

Exposure variables of interest included maternal characteristics (gravidity, level of education, and IPTp regimen received during pregnancy), infant characteristics (sex, sickle hemoglobin genotype, and whether or not the child was born small for gestational age), date the observation period began (before (Nov 21-May 23), during (Jun 23-Oct 23), or after (Nov 23 – Mar 24) the most recent universal LLIN distribution campaign), and household characteristics (household wealth and home construction). Principal component analysis was used to generate a wealth index based on ownership of common household items. Households were ranked by wealth scores and grouped into tertiles to give a categorical measure of socioeconomic status. House types were classified based on definitions previously developed for the study area ^15^. Modern houses were defined as having plaster or cement walls, metal or wooden roofs, and closed eaves; all other houses were defined as traditional.

Three outcome measures were assessed: (1) the incidence of symptomatic malaria, with an incident episode defined as the presence of fever (history of fever in the past 24 hours or a tympanic temperature ≥ 38.0 °C) with a positive thick blood smear not preceded by another malaria episode in the last 14 days; (2) the prevalence of microscopic parasitemia, defined as the proportion of routine samples positive for asexual parasites by microscopy; and (3) the prevalence of microscopic or submicroscopic parasitemia, defined as the proportion of routine samples positive for parasites by either microscopy or quantitative PCR.

For all analyses, a modified intention-to-treat approach was utilized, which included all children enrolled and followed through at least 12 weeks of age and without missing sickle cell status or household survey information. Time at risk was calculated as the time from 8 weeks of age and ended when the study participants reached 12 months of age or early study termination. Incident outcomes were compared using negative binomial regression models. Repeated prevalence measures were compared using log-binomial models with robust standard errors and generalized estimating equations to adjust for repeated measures in the same participant. Multivariate analyses included all exposure variables of interest assessed by univariate analyses (saturated models). In all analyses, 2-tailed *P* values < 0.05 were considered statistically significant.

### Ethical considerations

Ethical approval was obtained from the Makerere University School of Biomedical Sciences – Research and Ethics Committee -SBS-REC (SBS-2021-052), the Uganda National Council for Science and Technology (HS1819ES), and the Stanford University Institutional Review Board-IRB (62131). Written informed consent was obtained for all participants prior to enrollment into the cohort study.

## RESULTS

### Characteristics of study participants

Between November 2021 and March 2024, 912 infants were screened for eligibility, and 871 were enrolled into the study (**Figure 1**). Of the total enrolled participants, 855 were included in the final analysis after excluding 14 infants who were withdrawn before reaching 12 weeks of age, 1 who lacked sickle cell genotype results, and 1 who did not have a household survey done. Among those analyzed, 808 participants (94.5%) successfully completed follow-up through 12 months of age. A total of 41 participants were withdrawn prematurely, with the most common reason for withdrawal relocation outside the study area.

**Figure 1:**
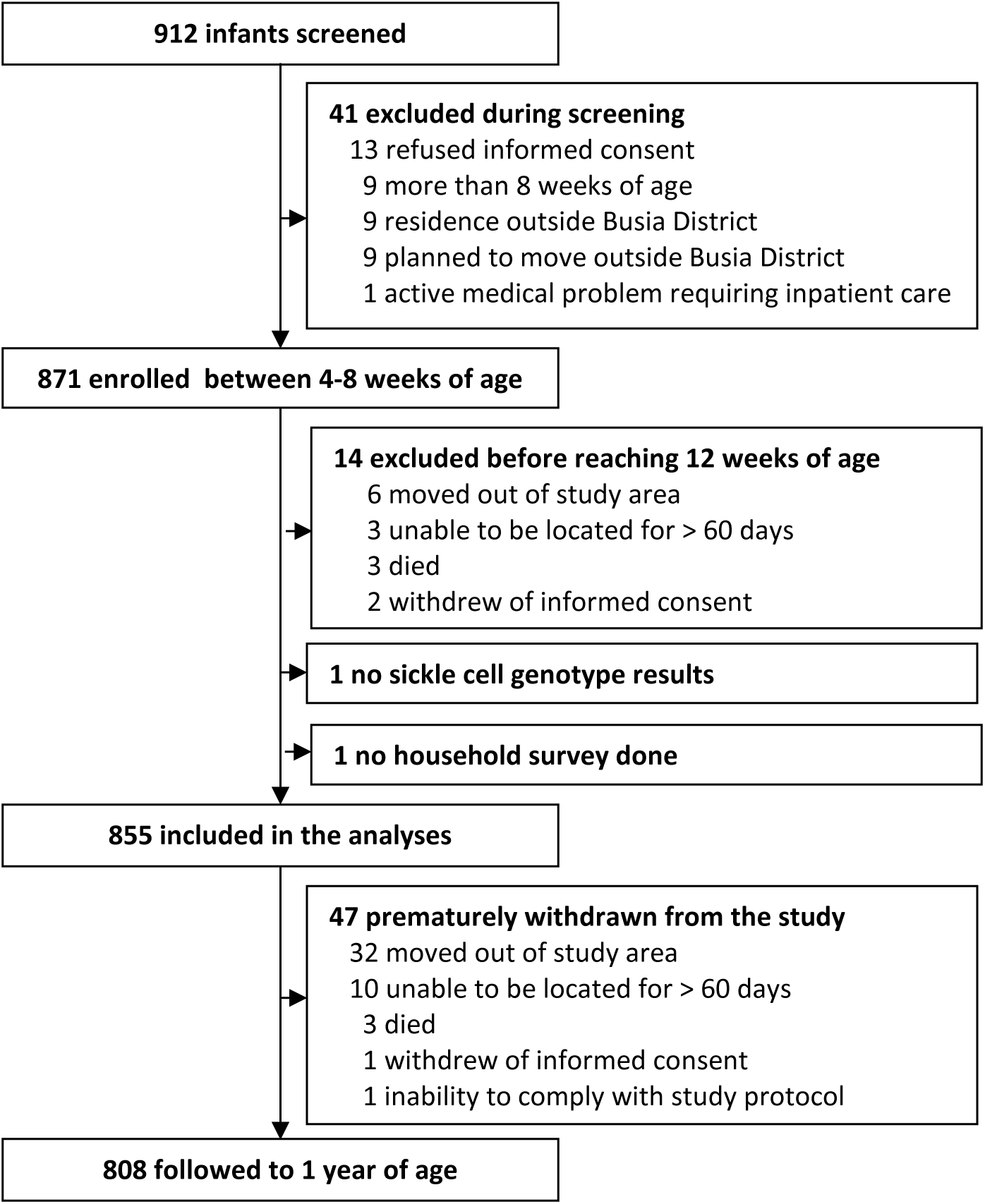
Study profiles.

Of the 855 infants included in the analyses, 23.2% were born to primigravid mothers, 69.6% were born to mothers with either primary education or no formal education, and 70.2% resided in traditional houses (**Table 1**). Overall, 48.4% were female, 21.2% had sickle cell trait, 0.7% had sickle cell disease, and 21.4% were born small-for-gestational age (SGA). The date the observation period began (8 weeks of age) occurred after universal LLIN distribution (October 2023) in 38.0% of infants (**Table 1**).

**Table 1.** Risk factors for the incidence of symptomatic malaria between 8 weeks and 1 year of age.

| **Risk factor** | **Category**<br>**(number of infants)** | **Episodes of malaria**<br>**(incidence PPY)** | **IRR (95% CI)** | **p-**<br>**value** | **aIRR (95% CI)** | **p-value** |
| --- | --- | --- | --- | --- | --- | --- |
| **Maternal and household characteristics** |  |  |  |  |  |  |
| Gravidity | Multigravid (657) | 500 (0.91) | Reference group |  | Reference group |  |
|  | Primigravid (198) | 162 (1.02) | 1.11 (0.83-1.47) | 0.48 | 1.14 (0.88-1.47) | 0.34 |
| Maternal Education | Secondary or higher (260) | 167 (0.78) | Reference group |  | Reference group |  |
|  | None or primary (595) | 495 (1.00) | 1.29 (0.99-1.68) | 0.06 | 1.34 (1.05-1.71) | 0.02 |
| Type of house construction | Modern (255) | 135 (0.65) | Reference group |  | Reference group |  |
|  | Traditional (600) | 527 (1.06) | 1.64 (1.24-2.16) | <0.001 | 1.60 (1.23-2.08) | <0.001 |
| Household wealth categories | Least poor (275) | 172 (0.76) | Reference group |  | Reference group |  |
|  | Middle (298) | 240 (0.97) | 1.28 (0.95-1.73) | 0.10 | 1.12 (0.85-1.48) | 0.42 |
|  | Poorest (282) | 250 (1.07) | 1.41 (1.04-1.91) | 0.03 | 1.04 (0.78-1.37) | 0.81 |
| Maternal IPTp regimen | SP (291) | 230 (0.96) | Reference group |  | Reference group |  |
|  | DP (290) | 235 (0.97) | 1.02 (0.76-1.36) | 0.90 | 1.13 (0.87-1.47) | 0.36 |
|  | DP + SP (274) | 197 (0.87) | 0.92 (0.68-1.23) | 0.56 | 1.05 (0.80-1.38) | 0.71 |
| **Infant characteristics** |  |  |  |  |  |  |
| Sex | Male (441) | 364 (1.00) | Reference group |  | Reference group |  |
|  | Female (414) | 298 (0.87) | 0.88 (0.69-1.12) | 0.30 | 0.97 (0.78-1.21) | 0.79 |
| SS genotype | AA (668) | 572 (1.03) | Reference group |  | Reference group |  |
|  | AS (181) | 85 (0.57) | 0.55 (0.40-0.76) | <0.001 | 0.61 (0.45-0.83) | 0.001 |
|  | SS (6) | 5 (1.05) | 1.04 (0.26-4.26) | 0.95 | 0.64 (0.19-2.18) | 0.48 |
| Born SGA | No (672) | 510 (0.92) | Reference group |  | Reference group |  |
|  | Yes (183) | 152 (1.00) | 1.09 (0.81-1.46) | 0.57 | 1.03 (0.79-1.34) | 0.83 |
| Date observation period began | Nov 21 – May 23 (340) | 471 (1.67) | Reference group |  | Reference group |  |
|  | Jun 23 – Oct 23 (190) | 101 (0.65) | 0.39 (0.29-0.52) | <0.001 | 0.38 (0.29-0.51) | <0.001 |
|  | Nov 23 – Mar 24 (325) | 90 (0.33) | 0.20 (0.15-0.26) | <0.001 | 0.20 (0.15-0.27) | <0.001 |

### Malaria burden over the follow-up period

Over the study period, there were 662 episodes of symptomatic malaria during 706.7 person-years of follow-up, corresponding to an overall incidence of 0.93 episodes per person-year (ppy). Incidence increased from 0.53 episodes ppy in children < 3 months of age to 0.86 in children age ≥3 - <6 months of age and 1.05 episodes ppy in children ≥6-12 months of age. The vast majority (650/662, 98.2%) of symptomatic malaria cases were uncomplicated. Among the 12 episodes of complicated malaria, 5 had only danger signs, 6 had severe anemia (hemoglobin < 5 gm/dL), and 1 had jaundice. There were no deaths due to malaria. Considering all 662 malaria episodes, 231 (34.9%) occurred prior to 6 months of age and 469 (70.8%) occurred prior to 9 months of age.

The overall prevalence of microscopic parasitemia at the time of routine visits was 7.9%, while combined microscopic and submicroscopic parasitemia prevalence was 21.8%. Parasitemia increased progressively with infant age: microscopic prevalence rose from 5.2% at 8–16 weeks to 8.9% by 32–40 weeks, while microscopic or sub-microscopic parasitemia rose from 13.6% to over 25% across the same age ranges.

A marked decline in both malaria incidence and parasite prevalence was observed following district-wide distribution of LLINs in October 2023. Malaria incidence dropped from 1.67 episodes per person-year during the pre-intervention period (November 2021 – October 2023) to 0.33 episodes per person-year during the post-intervention period (November 2023 – January 2025, **Figure 2**). Microscopic parasitemia fell from 12.8% in November 2021–October 2023 to 6.9% in November 2023–March 2024 and to 2.6% in April 2024–January 2025. Similar reductions were observed for microscopic or sub-microscopic parasitemia (34.7% to 17.2% to 9.8%).

**Figure 2:**
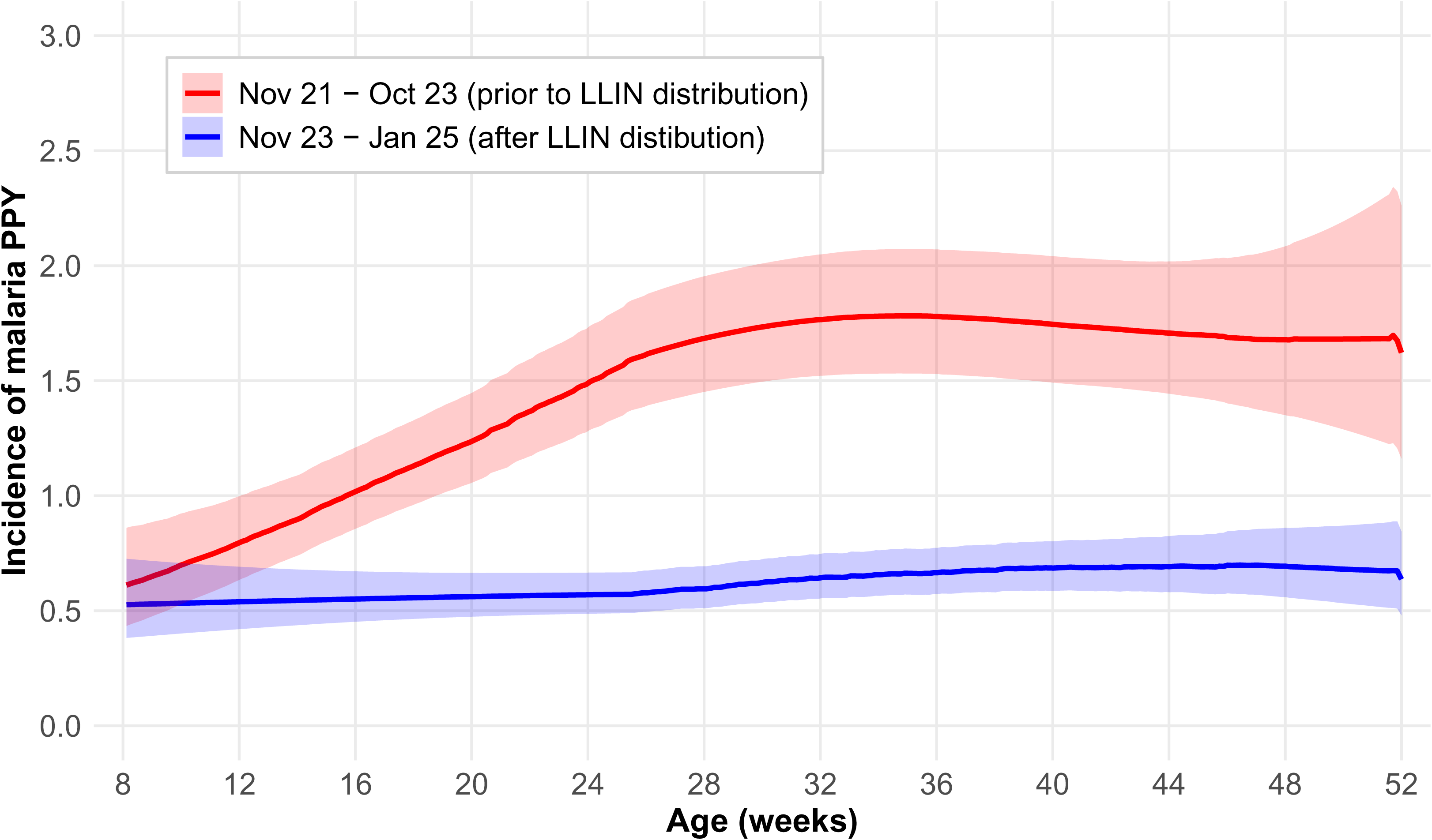
Incidence of malaria by age in weeks.

### Factors associated with the incidence of symptomatic malaria

To better understand factors associated with the incidence of symptomatic malaria, we first examined associations in univariate models (Table 1). Infants residing in traditional housing had a 64% higher incidence of symptomatic malaria compared to those in modern housing (1.06 vs. 0.65 episodes per person-year; IRR = 1.64, 95% CI: 1.24–2.16; p<0.001) and those living in the poorest tertile of households had a 41% higher incidence compared to those in the least poor houses (1.07 vs 0.76 episodes per person-year; IRR = 1.41, 95% CI: 1.04-1.91; p=0.03). A similar trend was seen for maternal education, with higher incidence among infants born to mothers with no or only primary education compared to those with secondary or higher (1.00 vs. 0.78 episodes per person-year; IRR = 1.29, 95% CI: 0.99–1.68; p=0.06). Infants with HbAS had 45% fewer episodes of malaria compared to those with HbAA (0.57 vs. 1.03 episodes per person-year; IRR = 0.55, 95% CI: 0.40–0.76; p<0.001). Regarding calendar time, incidence was highest among infants whose observation period began between November 2021–May 2023 (1.67 PPY), and then decreased to 0.65 PPY in those that began between June–October 2023 (IRR = 0.39; p<0.001) and to 0.33 PPY in those that began between November 2023–March 2024 (IRR = 0.20; p<0.001) (**Table 2**).

**Table 2.** Risk factors for microscopic parasitemia at the time of routine visits.

| Risk factor | Category | Parasite prevalence (%) | PR (95% CI) | p-value | aPR (95% CI) | p-value |
| --- | --- | --- | --- | --- | --- | --- |
| **Maternal and household characteristics** |  |  |  |  |  |  |
| Gravidity | Multigravid | 574/7715 (7.4%) | Reference group |  | Reference group |  |
|  | Primigravid | 206/2251 (9.2%) | 1.22 (0.93-1.60) | 0.15 | 1.22 (0.94-1.58) | 0.14 |
| Maternal Education | Secondary or higher | 204/3009 (6.8%) | Reference group |  | Reference group |  |
|  | None or primary | 576/6957 (8.3%) | 1.23 (0.93-1.63) | 0.14 | 1.27 (0.97-1.66) | 0.09 |
| Type of house construction | Modern | 131/2945 (4.5%) | Reference group |  | Reference group |  |
|  | Traditional | 649/7021 (9.2%) | 2.11 (1.53-2.91) | <0.001 | 1.92 (1.41-2.61) | <0.001 |
| Household wealth categories | Least poor | 183/3185 (5.8%) | Reference group |  | Reference group |  |
|  | Middle | 304/3483 (8.7%) | 1.51 (1.09-2.09) | 0.01 | 1.21 (0.88-1.65) | 0.24 |
|  | Poorest | 293/3298 (8.9%) | 1.55 (1.12-2.15) | 0.009 | 1.18 (0.85-1.63) | 0.33 |
| Maternal IPTp regimen | SP | 262/3385 (7.7%) | Reference group |  | Reference group |  |
|  | DP | 279/3408 (8.2%) | 1.07 (0.80-1.44) | 0.65 | 1.12 (0.85-1.48) | 0.43 |
|  | DP + SP | 239/3173 (7.5%) | 0.97 (0.72-1.31) | 0.85 | 1.08 (0.81-1.45) | 0.60 |
| **Infant characteristics** |  |  |  |  |  |  |
| Sex | Male | 406/5153 (7.9%) | Reference group |  | Reference group |  |
|  | Female | 374/4813 (7.8%) | 0.98 (0.77-1.26) | 0.90 | 1.06 (0.84-1.35) | 0.61 |
| SS genotype | AA | 596/7788 (7.7%) | Reference group |  | Reference group |  |
|  | AS | 180/2111 (8.5%) | 1.10 (0.82-1.49) | 0.51 | 1.14 (0.86-1.51) | 0.36 |
|  | SS | 4/67 (6.0%) | 0.83 (0.34-2.03) | 0.69 | 0.45 (0.16-1.28) | 0.13 |
| Born SGA | No | 569/7832 (7.3%) | Reference group |  | Reference group |  |
|  | Yes | 211/2134 (9.9%) | 1.37 (1.03-1.83) | 0.03 | 1.26 (0.96-1.66) | 0.10 |
| Age at time of routine visit | 8-16 weeks | 133/2537 (5.2%) | Reference group |  | Reference group |  |
|  | 20-28 weeks | 213/2515 (8.5%) | 1.61 (1.29-2.01) | <0.001 | 1.95 (1.55-2.44) | <0.001 |
|  | 32-40 weeks | 220/2479 (8.9%) | 1.68 (1.33-2.13) | <0.001 | 2.39 (1.87-3.05) | <0.001 |
|  | 44-52 weeks | 214/2435 (8.8%) | 1.67 (1.31-2.15) | <0.001 | 2.71 (2.09-3.51) | <0.001 |
| Date of observation | Nov 21 – Oct 23 | 536/4174 (12.8%) | Reference group |  | Reference group |  |
|  | Nov 23 – Mar 24 | 148/2145 (6.9%) | 0.73 (0.57-0.93) | 0.01 | 0.64 (0.50-0.82) | <0.001 |
|  | Apr 24 – Jan 25 | 96/3647 (2.6%) | 0.38 (0.29-0.50) | <0.001 | 0.21 (0.16-0.29) | <0.001 |

In multivariate models, both housing and maternal education remained independently associated with the incidence of symptomatic malaria. Infants in traditional housing had a 60% higher incidence of malaria (aIRR = 1.60; 95% CI: 1.23–2.08; p<0.001), and infants born to mothers with no or only primary education had a 34% higher incidence compared to those whose mothers attained at least secondary education (aIRR = 1.34; 95% CI: 1.05–1.71; p=0.02). The protective effect of sickle cell trait persisted, with HbAS associated with a 39% lower incidence of malaria (aIRR = 0.61; 95% CI: 0.45–0.83; p=0.001). The protective effect of calendar time in relationship to universal LLIN distribution was also robust, with 62% lower incidence in those whose observation period began between June–October 2023 and 80% lower incidence in those whose observation period began between November 2023–March 2024, compared to those whose observation period began between November 2021–May 2023. Other factors, including maternal IPTp regimen in the DPSP study, gravidity, infant sex, household wealth, and being born SGA, were not independently associated with the incidence of symptomatic malaria (**Table 1**).

### Risk factors for parasitemia during routine visits

Associations between exposures of interest and the risk of routine parasitemia largely mirrored those for symptomatic malaria, though there were some notable differences (**Tables 2 and 3**). In univariate analyses, the prevalence of microscopic parasitemia was significantly higher in children living in traditional compared to modern housing (9.2% vs. 4.5%; PR = 2.11; 95% CI: 1.53–2.91; p<0.001) and those living in the poorest houses compared to the least poor houses (8.9% vs. 5.8%, PR = 1.55; 95% CI: 1.12-2.15; p=0.009). Combined microscopic and submicroscopic parasitemia followed the same patterns (24.3% vs. 15.8%; PR = 1.56; 95% CI: 1.29–1.90; p<0.001 and 24.3% vs. 16.9%; PR = 1.45; 95% CI: 1.17-1.79; p=0.001, respectively). Maternal education followed a similar trend, with non-significantly higher prevalence among infants of less-educated mothers (microscopic PR = 1.23; p=0.14; combined PR = 1.18; p=0.07).

**Table 3.** Risk factors for microscopic or sub-microscopic parasitemia at the time of routine visits.

| Risk factor | Category | Parasite prevalence (%) | PR (95% CI) | p-value | aPR (95% CI) | p-value |
| --- | --- | --- | --- | --- | --- | --- |
| **Maternal and household characteristics** |  |  |  |  |  |  |
| Gravidity | Multigravid | 1653/7715 (21.4%) | Reference group |  | Reference group |  |
|  | Primigravid | 520/2251 (23.1%) | 1.07 (0.88-1.29) | 0.51 | 1.08 (0.92-1.27) | 0.37 |
| Maternal Education | Secondary or higher | 588/3009 (19.5%) | Reference group |  | Reference group |  |
|  | None or primary | 1585/6957 (22.8%) | 1.18 (0.99-1.42) | 0.07 | 1.18 (1.00-1.38) | 0.05 |
| Type of house construction | Modern | 466/2945 (15.8%) | Reference group |  | Reference group |  |
|  | Traditional | 1707/7021 (24.3%) | 1.56 (1.29-1.90) | <0.001 | 1.47 (1.23-1.76) | <0.001 |
| Household wealth categories | Least poor | 538/3185 (16.9%) | Reference group |  | Reference group |  |
|  | Middle | 835/3483 (24.0%) | 1.41 (1.15-1.74) | 0.001 | 1.18 (0.98-1.43) | 0.09 |
|  | Poorest | 800/3298 (24.3%) | 1.45 (1.17-1.79) | 0.001 | 1.18 (0.97-1.43) | 0.10 |
| Maternal IPTp regimen | SP | 733/3385 (21.7%) | Reference group |  | Reference group |  |
|  | DP | 771/3408 (22.6%) | 1.05 (0.86-1.28) | 0.62 | 1.07 (0.90-1.27) | 0.45 |
|  | DP + SP | 669/3173 (21.1%) | 0.97 (0.80-1.19) | 0.79 | 1.03 (0.86-1.23) | 0.73 |
| **Infant characteristics** |  |  |  |  |  |  |
| Sex | Male | 1128/5153 (21.9%) | Reference group |  | Reference group |  |
|  | Female | 1045/4813 (21.7%) | 0.99 (0.84-1.17) | 0.95 | 1.08 (0.94-1.24) | 0.30 |
| SS genotype | AA | 1724/7788 (22.1%) | Reference group |  | Reference group |  |
|  | AS | 436/2111 (20.7%) | 0.93 (0.75-1.15) | 0.51 | 1.02 (0.85-1.23) | 0.80 |
|  | SS | 13/67 (19.4%) | 0.98 (0.45-2.11) | 0.96 | 0.67 (0.35-1.28) | 0.22 |
| Born SGA | No | 1656/7832 (21.1%) | Reference group |  | Reference group |  |
|  | Yes | 517/2134 (24.2%) | 1.15 (0.95-1.39) | 0.16 | 1.08 (0.91-1.27) | 0.37 |
| Age at time of routine visit | 8-16 weeks | 345/2537 (13.6%) | Reference group |  | Reference group |  |
|  | 20-28 weeks | 583/2515 (23.2%) | 1.71 (1.50-1.94) | <0.001 | 2.00 (1.76-2.27) | <0.001 |
|  | 32-40 weeks | 623/2479 (25.1%) | 1.85 (1.59-2.13) | <0.001 | 2.49 (2.15-2.88) | <0.001 |
|  | 44-52 weeks | 622/2435 (25.5%) | 1.88 (1.62-2.19) | <0.001 | 2.80 (2.41-3.26) | <0.001 |
| Date of observation | Nov 21 – Oct 23 | 1447/4174 (34.7%) | Reference group |  | Reference group |  |
|  | Nov 23 – Mar 24 | 368/2145 (17.2%) | 0.76 (0.65-0.88) | <0.001 | 0.64 (0.53-0.71) | <0.001 |
|  | Apr 24 – Jan 25 | 358/3647 (9.8%) | 0.56 (0.47-0.66) | <0.001 | 0.28 (0.23-0.34) | <0.001 |

In multivariate analyses, housing and infant age remained independent predictors of parasitemia (Tables 2–3). Traditional housing was associated with a 92% higher risk of microscopic parasitemia (aPR = 1.92; 95% CI: 1.41–2.61; p<0.001) and a 47% higher risk of combined microscopic or submicroscopic parasitemia (aPR = 1.47; 95% CI: 1.23–1.76; p<0.001). In infants aged 44–52 weeks, the risk of microscopic parasitemia was nearly three-fold higher (aPR = 2.71; 95% CI: 2.09–3.51; p<0.001), and the risk of combined microscopic or submicroscopic parasitemia was almost three-fold higher (aPR = 2.80; 95% CI: 2.41–3.26; p<0.001) compared to that at 8–16 weeks of age. Compared to the period between November 2021–October 2023, risk of microscopic parasitemia declined by 36% between November 2023–March 2024 (aPR = 0.64; 95% CI: 0.50–0.82; p<0.001) and by 79% between April 2024–January 2025 (aPR = 0.21; 95% CI: 0.16–0.29; p<0.001). Reductions in combined microscopic and submicroscopic parasitemia were similar (36% and 72%, respectively).

Interestingly, unlike its strong protective association with symptomatic malaria, sickle cell trait was not associated with reduced parasite prevalence in both unadjusted and adjusted analyses. In adjusted analyses, HbAS infants had a slightly higher, though non-significant, risk of both microscopic (aPR = 1.14; 95% CI: 0.86–1.51; p=0.36) and combined microscopic and sub-microscopic parasitemia (aPR = 1.02; 95% CI: 0.85–1.23; p=0.80) compared to HbAA. This contrast suggests that HbAS does not decrease the risk of infection with malaria parasites, but rather confers protection against progression to symptomatic disease. Other covariates, including maternal IPTp regimen, gravidity, maternal education, household wealth, gender, and SGA status were not independently associated with parasitemia in multivariate analyses.

## DISCUSSION

This prospective cohort study provides contemporary evidence on the natural history and burden of malaria in infants during their first year of life in a high-transmission setting. Malaria represented a substantial health burden even in very young infants, with important implications for prevention strategies and clinical practice. The overall malaria incidence of nearly one episode per child per year observed in our cohort underscores the significant disease burden in this vulnerable population.

Limited literature exists on malaria prevention and management in young infants living in high, perennial transmission settings, contributing to continued lack of attention to this risk group in research and policy ^7, 16, 17^. Although the WHO recommends PMC with SP in settings with perennial transmission, it is not currently used in Uganda, where there are very high rates of SP resistance^18^, or in most other countries in sub-Saharan Africa. Malaria vaccination is being rolled out in Uganda and across Africa widely this year, but standard vaccine dosing begins at 5-6 months of age, and optimal vaccine efficacy is not reached until receipt of 3 doses (at 9 months of age following Uganda dosing guidelines). In our study, 35% of malaria episodes during the first year of life occurred prior to 6 months of age, and 71% occurred prior to 9 months of age.

Although most malaria cases in our cohort were uncomplicated, participants were encouraged to come to a dedicated study clinic open 7 days per week for all medical care and provided with transportation reimbursement. In a real world setting, where access to prompt and effective care may be lacking, the burden of severe disease would likely be higher, especially in the youngest children due to immature immunity ^19^. There is an urgent need for the development and testing of age-appropriate preventive interventions targeting young infants, such as PMC with DP or other drugs, malaria vaccines given at earlier ages, administration of protective monoclonal antibodies^20^, and/or permethrin-treated baby wraps ^21^. Furthermore, treatment and chemoprevention guidelines often lack clear diagnostic and dosing recommendations for young infants and those weighing less than 5 kg^22^. This indicates a need for tailored diagnostic protocols and drug formulations that are palatable and weight specific to this age group. Addressing these gaps will be essential to reduce early childhood morbidity and mortality from malaria.

Sickle cell trait was relatively common in this high transmission setting, consistent with published estimates from Uganda^23^, and the differential effects of sickle cell trait on symptomatic malaria versus parasitemia provide insights into the mechanisms of protection. While HbAS conferred significant protection against symptomatic malaria (39% lower incidence), consistent with prior studies ^8^ ^24^, it showed no protective effect against parasitemia (both microscopic and submicroscopic). This pattern suggests that sickle cell trait does not decrease the risk of initial infection, but rather protects against progression to clinical disease, consistent with proposed mechanisms involving reduced parasite multiplication or enhanced immune recognition of infected cells ^25,26^.

The dramatic reduction in malaria incidence following district-wide distribution of LLINs containing alphacypermethrin plus chlorfenapyr in October 2023 provides strong evidence for the effectiveness of next-generation, dual active ingredient bednets in protecting infants. The 80% reduction in incidence (from 1.67 to 0.33 episodes per person-year) demonstrated that LLINs with alphacypermethrin plus chlorfenapyr can provide substantial protection even in areas with pyrethroid-resistant mosquitoes. This finding supports WHO recommendations for the use of next-generation LLINs in areas with confirmed pyrethroid resistance. The parallel reductions in both microscopic and submicroscopic parasitemia prevalence following LLIN distribution suggest that the intervention reduced malaria transmission and disease^27, 28^. This has broad implications for community-level malaria control, as reduced transmission can benefit both protected and unprotected individuals through indirect effects ^29^. In addition, these results emphasize the importance of maintaining regular distribution campaigns and ensuring proper and consistent net usage at the household level to sustain and further the gains made in malaria control and preventing malaria in infancy.

Our findings also highlight the persistent influence of socioeconomic factors on malaria risk in infants. Infants residing in traditionally constructed houses—typically characterized by mud walls, thatched roofs, and open eaves—are at a higher risk of exposure to malaria vectors ^30, 31^. Traditional housing construction was the strongest predictor of both symptomatic malaria and parasitemia, with infants in traditional houses experiencing 60% higher incidence of clinical episodes and nearly twice the risk of microscopic parasitemia. In contrast, improved housing structures with sealed walls and screened windows offer a protective barrier. The association between maternal education and malaria risk, while modest, underscores the importance of health literacy and decision-making capacity in malaria prevention. Mothers with higher education may be more likely to adopt preventive measures such as consistent use of insecticide-treated nets^32^, and seeking prompt medical care which translate into reduced malaria infection rates ^32, 33^.

Interestingly, in this study, maternal characteristics associated with protection against malaria in pregnancy – gravidity and receipt of more effective IPTp regimens – were not associated with malaria risk in infants. This contrasts with results from several observational studies which have suggested that malaria during pregnancy, particularly placental malaria, may increase the risk of infant malaria ^34, 35, 36^. However, this association is likely confounded by shared exposure risks between mother and infant. Our data suggest that mechanisms of protection during pregnancy may not impact infant outcomes.

Our study had several strengths, including its large sample size, prospective design, high level of retention, and use of both microscopy and qPCR to measure malaria burden. However, there were some limitations. First, the study was conducted in a single geographic region with specific transmission dynamics and healthcare practices, which may limit the generalizability of the findings to other malaria-endemic settings with different environmental, socio-economic, or healthcare factors. Second, while the decline in malaria prevalence following LLIN distribution supports the effectiveness of this intervention, the study lacked detailed individual-level data on net use and maintenance, which would have strengthened our causal inference. Third, the number of infants with sickle cell disease was small, limiting the power to study its association with malaria outcomes.

This study demonstrated that malaria represents a substantial and underappreciated burden in infants during their first year of life in high-transmission settings. The early onset of susceptibility, marked biological and social determinants of risk, and dramatic impact of vector control interventions have important implications for prevention strategies and clinical care.

Addressing this burden will require expanded prevention strategies targeting younger infants, continued investment in effective vector control, and attention to the social determinants that perpetuate malaria risk in the most vulnerable populations. The evidence presented here should inform policy discussions about extending malaria prevention strategies to cover the critical first months of life and guide clinical practice toward greater recognition of malaria risk in young infants.

## Data Availability

The datasets reported herein will be made available from the corresponding author on reasonable request.

## ACKNOWLEDGEMENTS

We would like to thank study participants for willingly accepting to give their consent and to participate in the study, study team at Masafu Hospital and Tororo Hospital and the administration of the Infectious Diseases Research Collaboration, and Stanford University for all their contributions.

